# Cell-free DNA methylome and fragmentome analysis for disease relapse monitoring in patients with Ewing Sarcoma

**DOI:** 10.1101/2025.04.25.25325254

**Authors:** Sophie A. Richardson, Gayatri Gulati, Patrick J.B. Harker, Alan Redfern, Simon P. Pearce, Mina Karimpour, Steven M. Hill, Vsevolod J. Makeev, Bernadette Brennan, Alexander TJ Lee, Alexandra Clipson, Caroline Dive, Dominic G. Rothwell, Martin G McCabe, Florent Mouliere

## Abstract

Liquid biopsies and cell-free DNA (cfDNA) offer minimally invasive methods for the diagnosis and monitoring of Ewing Sarcoma (EwS). EwS have a low tumour mutational burden and their detection with plasma cfDNA is challenging.

We hypothesised that analysing the cfDNA methylome and fragmentome could enhance sensitivity for detecting EwS and identifying early disease recurrence. We conducted whole-genome and methylome sequencing of cfDNA from 68 serial samples of 15 patients with EwS and 3 patients with CIC-rearranged sarcoma (CIC). With EwingSign, a new machine learning model, we identified EwS or CIC in a test set for 10 out of 12 patients at diagnosis and 15 out of 18 clinically confirmed relapse events. 0 out of 29 non-cancer controls were detected positive with EwingSign.

These findings indicate that cfDNA methylome and fragmentome analysis, if validated in a larger cohort, could improve disease detection, monitoring and relapse identification in patients with EwS.

## 1 Introduction

Ewing sarcoma (EwS) is the second most common bone and soft tissue cancer in children and young adults, with an incidence of 1 case per 1.5 million globally^1^. EwS is driven by a fusion between FET and ETS family genes, most commonly EWSR1::FLI1^2^. Disease relapse occurs in approximately 30-40% of patients with localised EwS and 60-80% of patients with metastatic disease^3^.Patients with EwS have a worse prognosis when relapse occurs within the first 2 years after diagnosis^3^, therefore monitoring of disease throughout this period is key to detecting relapse early. The standard of care for EwS relapse detection includes lung screening with chest X-ray and either CT or MRI scans to image the primary tumour site^4^. These methods are limited by their sensitivity for detecting small volume tumours and asymptomatic relapse events, and by the specialised equipment and staff required. Earlier detection and treatment of relapse when active disease bulk is minimal could lead to better survival outcomes. There is a clear, unmet need for a biomarker to detect relapse earlier in patients with EwS in a readily implemented and cost-effective manner.

Liquid biopsies are bio-fluids collected from the body, most commonly blood, and therefore are minimally invasive^5^. They contain cell-free DNA (cfDNA), RNA and other analytes released into the bloodstream^6^. Cancer cells also release DNA into the bloodstream, referred to as circulating tumour DNA (ctDNA)^7^. Hence, the detection of both genetic and epigenetic features of cancer is possible with a liquid biopsy^8^. In the context of EwS, previous studies have detected the EWSR1::FLI1 fusion gene in cfDNA^9–14^ or utilised the fragmentation patterns of cfDNA to detect EwS signal in the blood of patients^15^. The EWSR1::FLI1 fusion^9,14^, copy number aberrations (CNAs)^15–17^ and cfDNA fragmentation^15,17^ have been monitored throughout diagnosis and relapse in a cohort of up to 95 patients with EWS. cfDNA methylation has been explored in liquid biopsies for diagnosis of EwS^18,19^, with evidence of distinct epigenetic features of EwS compared to other sarcomas apparent through tumour tissue analysis^20,21^ as well as interpatient heterogeneity within EwS^22^. cfDNA methylation has also been shown to improve sensitivity of cancer detection compared to mutation-based analysis^23^ and has the capability to determine tissue of origin^24,25^. This study aims to longitudinally profile cfDNA samples from patients with EwS and CIC-rearranged sarcoma (CIC) using the in-house T7-MBD-seq assay, an enrichment-based methylation capture method^25,26^. T7-MBD-seq is an integrated workflow combining genome wide methylation analysis and shallow whole genome sequencing (WGS) for the recovery of CNAs and cfDNA fragmentomic features. A machine learning classifier, which we termed EwingSign, was developed to detect EwS or CIC using cfDNA methylation features. Longitudinal analysis was performed to understand how ctDNA burden, cfDNA fragmentation and cfDNA methylation changed over time, following systemic treatment and with sequential episodes of disease relapse. Finally, all individual features of the assay were compared to determine their performance for detection of EwS- and CIC-associated signal at both diagnosis and relapse.

## 2 Results

### 2.1 Study Cohort and Design

In total 70 plasma samples were collected from 18 patients with sarcomas: 15 with EwS and 3 with CIC (**Figure 1**). For 12 patients, diagnosis samples were collected prior to treatment, (11 EwS and 1 CIC). For 10 of these patients, two plasma samples were collected during first-line chemotherapy (20 samples). For 9 patients (6 EwS, 3 CIC), plasma samples were collected at one or more disease relapse timepoints (18 samples), and in 7 of these patients, liquid biopsies were also collected during treatment of relapsed disease (20 samples). The mean cfDNA concentration was 24.11 ng/mL of plasma (0.08 - 356.74 ng/mL of plasma, **Supplementary Figure 1**). Two first-line treatment samples were excluded from further analysis due to low cfDNA concentration (< 1 ng/mL of plasma). The remaining 68 cfDNA samples were analysed with T7-MBD-seq^26^. WGS was performed to a mean 1.66-fold coverage (0.5-4X). Additionally, 107 non-cancer controls (NCCs) were analysed with T7-MBD-seq and split into a training set (n = 78) and an unseen test set (n = 29, **Figure 1**). Of the NCCs, 98 were previously published^25,26^ and 9 are new. The proportion of ctDNA (ichorTF) was calculated based on the CNA profile of WGS (**Supplementary Figure 2**) with ichorCNA^27^. The proportion of short cfDNA fragments (p100-150 bp) was determined from the same WGS data^15,28^. Methylation array data from Koelsche et al.^21^ and the training NCC set were used to determine differentially methylated windows (DMRs) between EwS and NCC, CIC and NCC, and EwS and CIC (**Supplementary Figure 3**). The top 200 intersecting DMRs were selected for downstream analysis using the mean β-value (β mean), a scaled measure of methylation between 0 and 1, and EwingSign.

**Figure 1.**
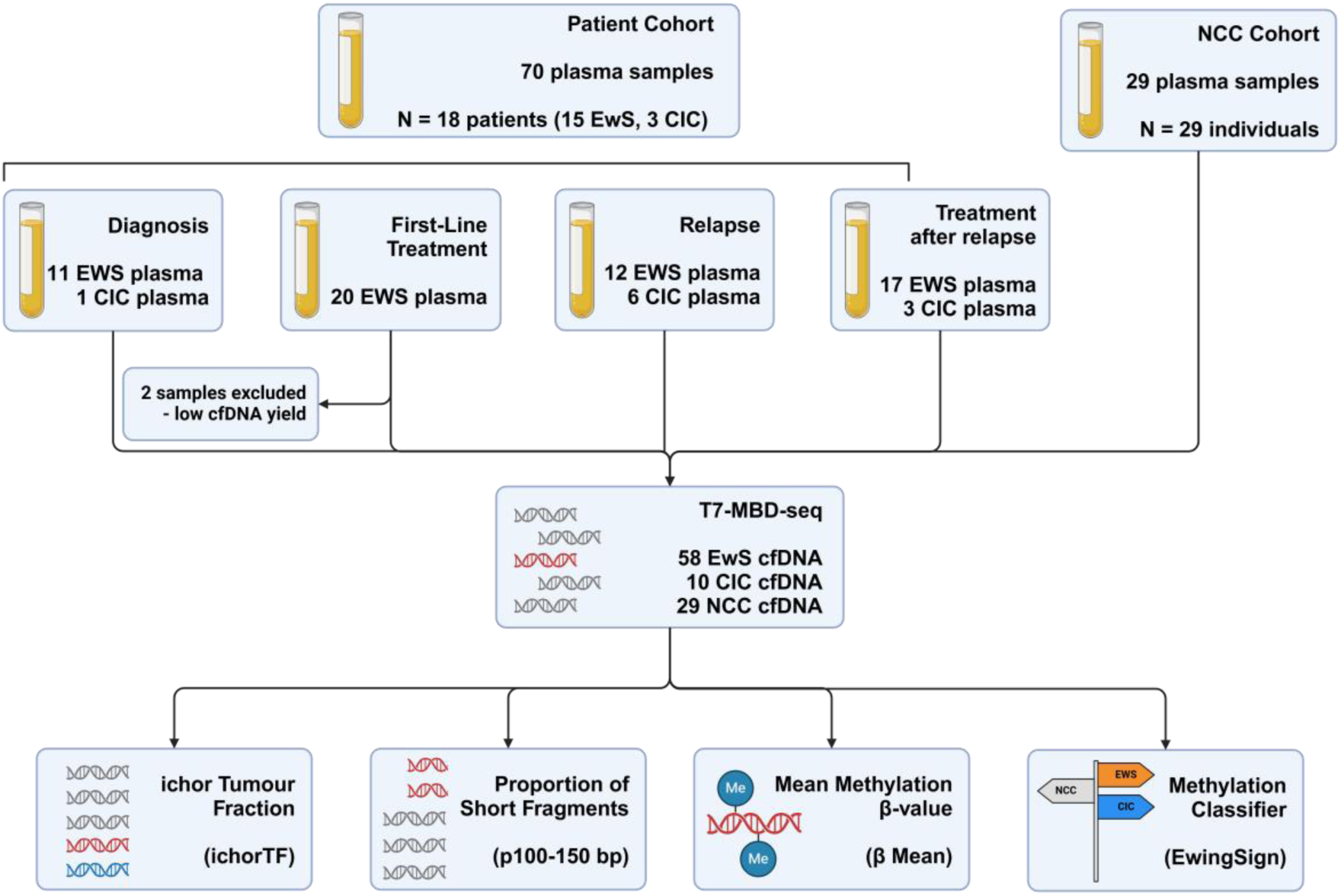
An overview of the study design. Study cohort flow chart depicting the number of patients and samples at each event, and the inclusion and exclusion of samples in cfDNA analysis. Analysis workflow shown also, describing the analysis performed (copy number, cfDNA fragmentation and cfDNA methylation), and the outputs of these analyses (ichorCNA Tumour Fraction, proportion of short fragments, mean methylation β-value and methylation classifier).

### 2.2 A methylation classifier, EwingSign, is the most sensitive and specific modality for detection of EwS and CIC disease using cfDNA

Using copy number aberrations, ctDNA was detectable in 6/12 diagnosis samples, 0/18 first-line treatment samples, 10/18 samples at disease relapse and 4/20 samples taken on treatment after relapse (cut-off of 0.03 ichorTF, **Figure 2A**). None of the 29 samples from NCCs were positive for ctDNA using the ichorTF method. The ichorTF was significantly higher in samples from relapse events compared to NCCs (p < 0.05, Wilcoxon test). However, there was no significant difference in ichorTF between patients at diagnosis and NCCs (p > 0.05, Wilcoxon test).

**Figure 2.**
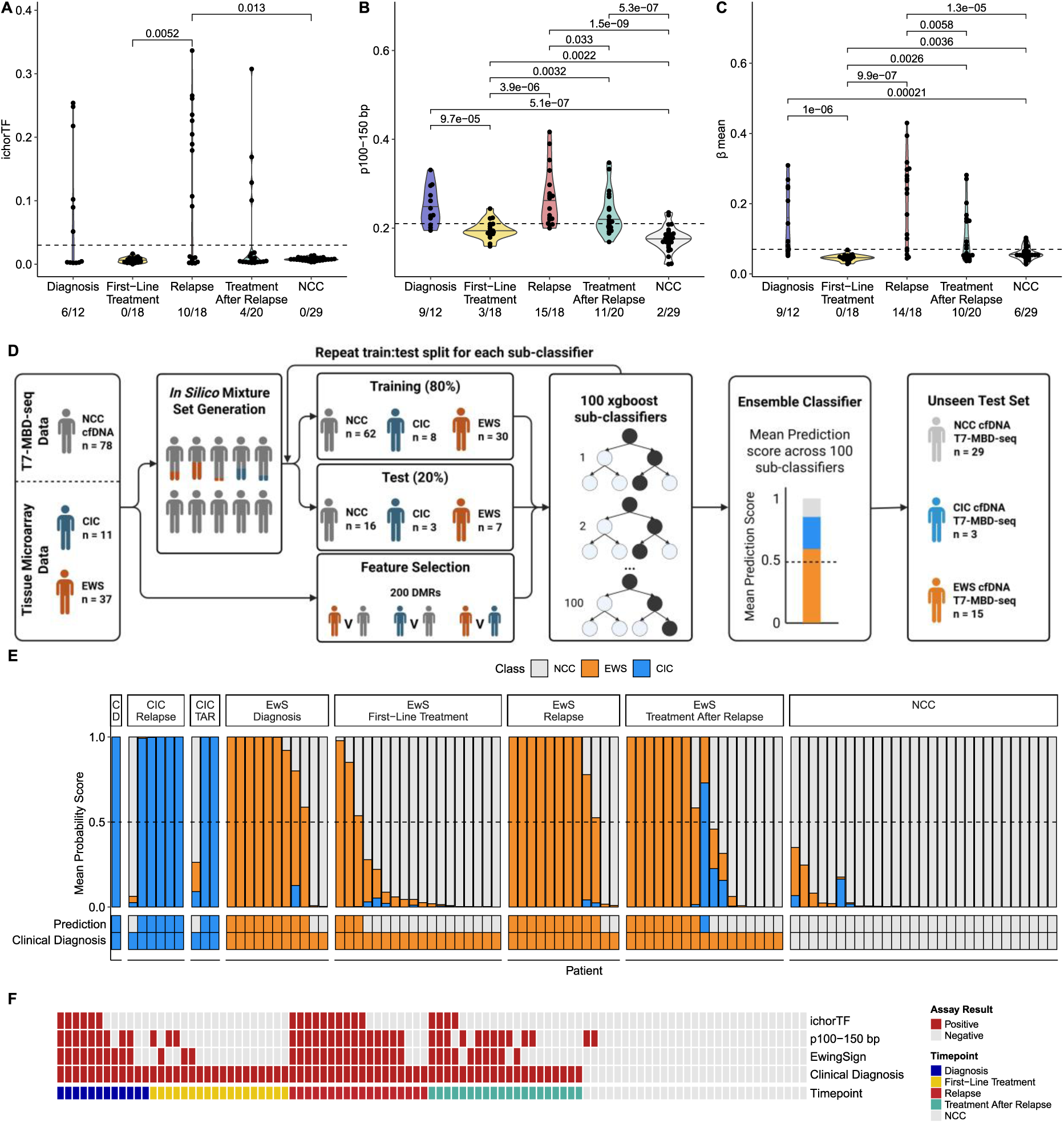
A methylation classifier, EwingSign, is the most sensitive and specific modality for detection of EwS and CIC disease. **A-C:** Violin plots of ichorCNA tumour fraction (**A**), Proportion of short fragments (100-150 bp) (**B**) and mean methylation beta value across 200 differentially methylated regions (DMRs) (**C**) at diagnosis, first-line treatment, relapse, treatment after relapse and in non-cancer controls. Wilcoxon test used for comparisons, only significant p values shown (p < 0.05). **D:** Graphical description of the EwingSign classifier. NCC cfDNA and tumour tissue microarray data was used for feature selection and generation of in silico mixture sets. These features and in silico mixtures from 80% of the input data were used to train an xgboost classifier, then the remaining in silico mixtures from 20% of the input data were used to test the classifier. This process was repeated until 100 xgboost sub-classifiers were generated. An ensemble of the 100 classifiers was obtained using the mean scores across all 100 classifiers. The ensemble classifier was then applied to an unseen test set of NCC, CIC and EWS cfDNA. **E:** EwingSign classifier results. Each bar indicates the results of an individual sample. The proportion of the bar fill colour indicates the EwS (orange), CIC (blue) and NCC (grey) scores, which all sum to a value of 1. Samples are faceted by cancer type first (CIC, EwS and NCC), then by timepoint (diagnosis, first-line treatment, relapse and treatment after relapse). Facet labels are abbreviated where necessary: C = CIC, D = Diagnosis, TAR = Treatment after relapse. **F:** A heatmap showing the frequency of positive (red) and negative (grey) assay results for each modality. From top to bottom, the rows show: ichorTF, p100-150 bp, EwingSign, Clinical Diagnosis. The final row indicates whether the sample was taken at diagnosis (blue), first-line treatment (yellow), relapse (red) or treatment after relapse (green) for the patient samples, or from a NCC (grey).

Size distribution of cfDNA was also calculated from the WGS data (**Supplementary Figure 4**). Based on the difference between the NCC and patients with EwS and CIC, and a previous publication from Peneder et al.^15^, we chose the proportion of fragments between 100-150 bp (p100-150 bp) to represent tumour-associated cfDNA. p100-150 bp was deemed to be positive if the value exceeded the threshold of 0.21. 9/12 diagnosis samples, 3/18 on treatment samples, 15/18 relapse samples and 11/20 samples taken on treatment after relapse were positive with 2/29 NCCs also above this threshold (**Figure 2B**). In diagnosis samples, the p100-150 bp was significantly greater than first-line treatment samples and NCC samples (p < 0.05, Wilcoxon test). The p100-150 bp was significantly greater in relapse samples compared to samples taken at all other timepoints except for diagnosis and in NCC samples (p < 0.05, Wilcoxon test). The p100-150 bp was also significantly greater in samples taken at either first-line treatment or treatment after relapse and NCC samples (p < 0.05, Wilcoxon test).

Using the methylation enrichment component of T7-MBD-seq, the β mean was determined for each sample. The β mean was calculated across the top 200 DMRs described above. The β mean was positive if the value exceeded a threshold of 0.07. Using the β mean, 9/12 diagnosis samples, 0/18 on treatment samples, 14/18 relapse events and 10/20 samples taken on treatment after relapse were positive (**Figure 2C**); 6/29 NCCs were also positive. The β mean was significantly greater in diagnosis samples compared to first-line treatment samples and NCC samples (p < 0.05, Wilcoxon test). In relapse samples, the β mean was significantly greater in NCC samples and in samples taken at all timepoints except diagnosis (p < 0.05, Wilcoxon test). Contrary to the p100-150 bp, a significantly lower β mean was observed in the first-line treatment samples compared to the NCC samples (p < 0.05, Wilcoxon test), but there was no significant difference between the samples taken at treatment after relapse and the NCC samples (p > 0.05, Wilcoxon test).

Next, the same 200 DMRs were used as features in our machine learning model, EwingSign, to classify samples as either EwS, CIC or NCCs (**Figure 2D**). Briefly, an augmented dataset was generated with *in silico* mixed samples of EwS tissue or CIC tissue methylation array data published by Koelsche et al.^21^ with the NCC T7-MBD-seq training set data with a uniform distribution of tumour fractions (0.005-0.1). The data from EwS and CIC tumour tissue and the NCC training set were then split into an 80:20 training to test data split. Each of the 100 sub-classifiers used a different 80:20 split of the tissue and NCC training set data. The *in-silico* mixed samples were used to train and test the classifier prior to the application of EwingSign, to the unseen test set of EwS (15 patients, 58 samples), CIC (3 patients, 10 samples) and NCC (29 NCCs, 29 samples) cfDNA T7-MBD-seq data. EwingSign reports the mean prediction score for EwS, CIC or NCC class based on an ensemble of the 100 sub-classifiers (**Supplementary Figure 5**). A sample was deemed to be classified if a single class exceeded 0.5 using the ensemble classifier. Using this methodology, 10/12 diagnosis samples, 3/18 first-line treatment samples, 15/18 relapse samples and 8/20 samples taken on treatment after relapse were correctly classified as either belonging to the EWS or CIC class (**Figure 2E**). One EwS sample taken on treatment after relapse was incorrectly classified as CIC, however this classifier is not intended to be a diagnostic tool so this is permissible in this context. All NCCs were correctly classified as NCC.

In summary, using EwingSign alone resulted in the greatest sensitivity (83.3%, **Supplementary Table 1**) for detection of disease in this cohort of patients compared to any of the other modalities, or in combination with p100-150 bp and/or ichorTF with 10/12 diagnosis (untreated) samples and 15/18 samples at the timepoint of disease relapse determined to be positive (**Figure 2F**). 0/29 NCCs were falsely positive using EwingSign, giving a specificity of 100%. The addition of ichorTF did not increase sensitivity or specificity, and addition of p100-150 bp decreased specificity and did not increase sensitivity.

### 2.3 Longitudinal liquid biopsy monitoring of EwS patients reflects disease dynamics

To detect relapse in a clinical setting, patients would be monitored longitudinally for disease with imaging techniques. We propose that liquid biopsies could augment this approach providing additional molecular information. Here, we report the analysis of longitudinal blood samples from four exemplar patients with different disease trajectories and clinical outcomes (**Figure 3A-D**).

**Figure 3.**
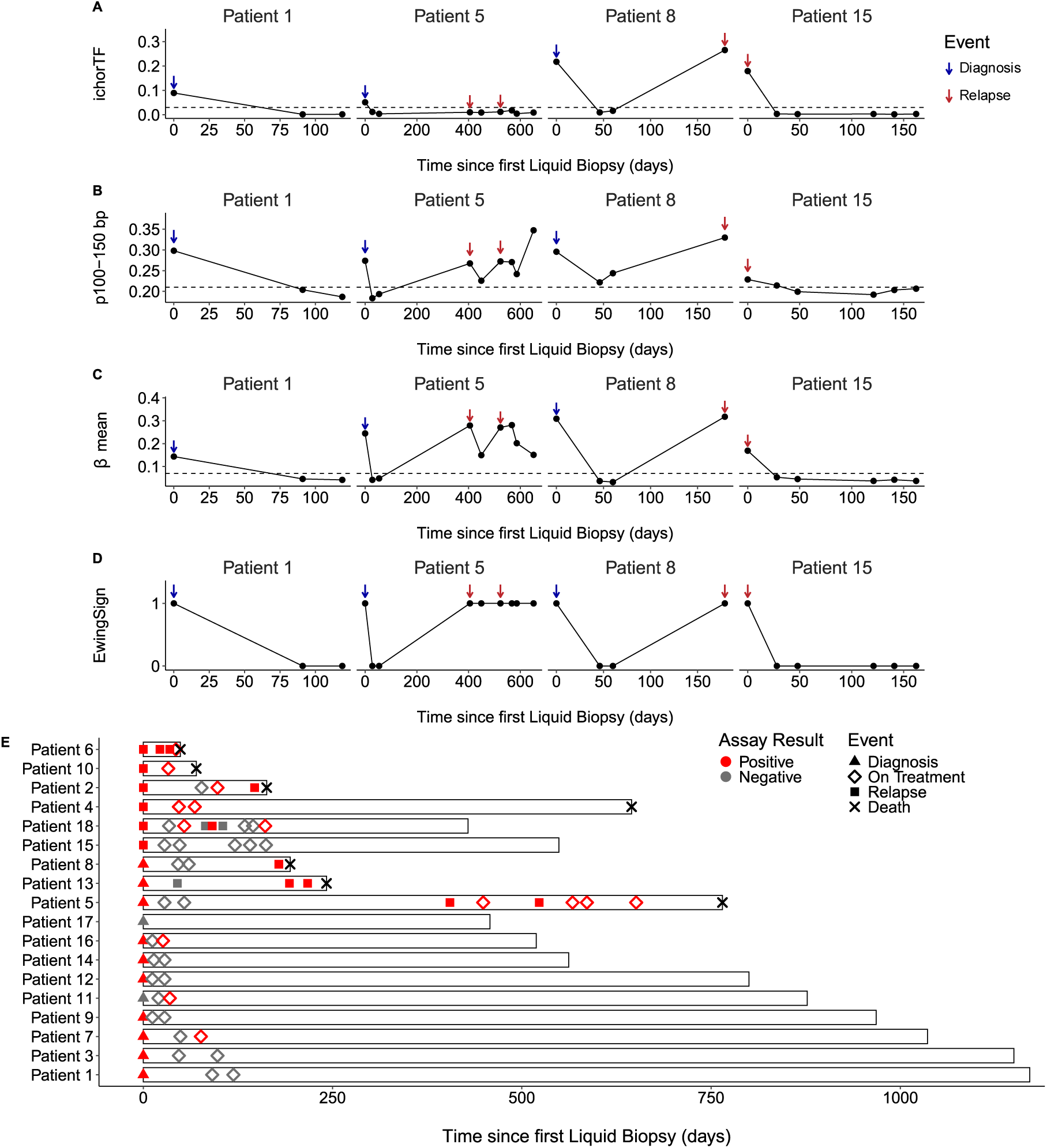
Longitudinal liquid biopsy monitoring of patients with EwS reflects disease dynamics. **A-D:** Longitudinal monitoring over time (days) using ichorTF (**A**), p100-150 bp (**B**), β mean (**C**) and EwingSign (1 = positive for EwS, 0 = negative for EwS, **D**) for patients 1, 5, 8 and 15. Blue arrows indicate diagnosis events, red arrows indicate relapse events, and an absence of arrows indicates an on-treatment event. Each point indicates a liquid biopsy sample taken, and its result. **E:** Swimmer’s plot of all patients in the cohort (n = 18). Bar length indicates the length of time the patient has been monitored for, with 0 indicating the date of the first liquid biopsy taken. The shape of the point indicates which event the sample was taken, either diagnosis (triangle), on treatment (diamond) or relapse (square). Death is shown with a black cross at the end of the bar, with absence of the cross indicating the patient is alive. The colour of the point indicates whether the assay result was positive (red) or negative (grey). A positive assay result is defined by a classification of EWS or CIC by EwingSign.

Patient 1 responded clinically to first-line treatment and did not relapse. Liquid biopsy samples at diagnosis were positive by all four modalities (**Figure 3A-D**), but all assay values were below the threshold for positivity at the two on-treatment timepoints at days 91 and 119.

Patient 5 relapsed after initial treatment, progressed rapidly through multiple subsequent lines of treatment and died. The sample was positive for all four modalities at diagnosis, then below assay thresholds and classified as NCC throughout first-line treatment (**Figure 3A-D**). At the first relapse timepoint (day 405), the sample was positive by p100-150 bp and β mean, and classified as EWS by EwingSign, but was negative by ichorTF. During treatment of this relapse and a second relapse, the samples continued to be positive for p100-150 bp and β mean, and classified by EwingSign as EWS, but ichorTF remained negative. Patient 5 then died 765 days after their diagnosis liquid biopsy.

Patient 8 did not respond to initial treatment and experienced a single relapse event. Diagnosis samples were positive by all four modalities (**Figure 3A-D**). First-line treatment samples were negative for ichorTF and β mean and classified as NCC by EwingSign. However, p100-150 bp was positive in first-line treatment samples. After an initial decrease, p100-150 bp increased from day 46 to day 60. A further increase was observed in all modalities from day 60 to the clinical diagnosis of relapse at day 179, at which point EwingSign also classified the sample as EwS. The patient died 194 days after their diagnosis liquid biopsy.

Patient 15 had successful treatment of relapsed disease. Samples at relapse were positive by all four modalities. Five subsequent on-treatment samples at days 28, 48, 121, 141 and 162 were all below assay thresholds, with no detection of EwS by EwingSign (**Figure 3A-D**). Despite monitoring for 596 days there was no evidence of progressive disease either by liquid biopsy or clinically.

Patients who experienced at least one relapse had positive liquid biopsy results in at least one relapse timepoint (9/9, **Figure 3E**, **Supplementary Figure 6**). These early data suggest these patients could have derived a benefit from liquid biopsies for detection of their relapse.

## 3 Discussion

We sought to determine a sensitive, high-throughput and cost-effective approach for detection of disease relapse of bone sarcomas (EwS and CIC) via multimodal analysis of cfDNA at baseline and in longitudinal blood samples using a single workflow, T7-MBD-seq. We show that using a cfDNA methylation machine learning classifier, EwingSign, provides a sensitive and specific detection of disease at both diagnosis and at disease relapse. Using EwingSign, all patients with relapsed disease had at least one positive relapse signal in their blood sample, with 83% of relapse events detected (15/18). At first relapse, 8/9 patients had a positive liquid biopsy result, suggesting that these patients could have benefited from monitoring with a cfDNA-based liquid biopsy.

Previous studies of liquid biopsies in EwS have focused on detection of the EWSR1::FLI1 fusion^9–14^, which is specific to EwS. Such targeted approaches may not be the most sensitive methods for detection of relapse in EwS patients. Therefore, we built upon the research of Peneder et al.^15^ to determine the utility of cfDNA fragmentation as a tool for monitoring EwS patients. As observed by Peneder et al.^15^, we also saw a high p100-150 bp fragments in a patient with no detectable ichorTF at either of their relapse timepoints. This was not unexpected as only 63% of EwS cases were found to have CNAs using tumour tissue^29^ and CNAs are a requirement for detection of ctDNA using this technique. In addition to high p100-150 bp fragment levels, patient 5 also had a high β mean and was classified as EwS by EwingSign, confirming that low ichorTF was associated with a lack of CNAs as opposed to lack of ctDNA. Furthermore, we also observed no significant difference between p100-150 bp fragmentation at diagnosis and relapse, as also seen by Peneder et al.^15^. Additionally, we explored the dynamics of each assay modality and observed an increase in tumour-associated signal upon relapse, which was also observed with ddPCR quantification of EWSR1::FLI1 fusion molecules^9^. As well as analysing the results of our EwingSign classifier longitudinally, we also observed the correct classification of 38/39 CIC and EwS samples where a cancer classification was made, with only one EwS sample incorrectly classified as CIC. This implies that the differential methylation of sarcomas in tissue^21^ may also be detectable in liquid biopsies using cfDNA methylation.

Whilst the results presented here are promising, the following limitations apply: a small cohort, irregular sampling schedule, retrospective analyses, a non-risk matched NCC cohort and limited genomic analyses. The small cohort size is due to the rarity of EwS (∼2% of teenage and young adult cancer cases^1^) and this being single centre study. For training EwingSign, we have mitigated the challenge of a limited training cohort by generating an augmented dataset using a cohort of tumour tissue data. Further validation in external cohorts will be required to validate our observations. The addition of further training and test data from tumour samples could also improve the classifier performance. The irregular, and opportunistic sampling schedule means we do not have the same sampling timepoints for every patient. Additionally, patients were not monitored with liquid biopsies after initial treatment and relapse samples were only collected when disease was detected by standard of care monitoring. Therefore, we could not explore whether liquid biopsies detect relapse earlier than current standard of care and this will be an essential next step. Of note, one patient was observed to have early indications of relapse using p100-150 bp fragment analysis (patient 8). This cohort was analysed retrospectively and, in the future, a clinical trial with a validated liquid biopsy assay for patient monitoring and decision-making would be required to determine whether earlier detection of relapse is possible, and to explore what the benefits of earlier detection of EwS relapse would be. Currently, there is no standard of care for treatment of relapsed EwS, however, there are clinical trials exploring various treatment options for treatment of patients with relapsed EwS, such as rEECur^30^. Additionally, our NCC cohort (median age = 62 years) was not age- or risk-matched, and is significantly older (p < 0.01, Wilcoxon test) than our patient cohort (median age = 22 years) due to the challenge of obtaining NCCs from a younger population. Future studies should take this into account and determine how the age of a NCC cohort impacts on cfDNA modalities.

Here, we used epigenetic features of cfDNA for detection of and patient monitoring for a EwS or CIC sarcoma signal. However, there is also scope for these cfDNA based assay features to be used to detect normal tissue toxicity and organ damage that may be caused by aggressive chemotherapy regimens used in paediatric patients with sarcoma^31,32^. cfDNA fragmentation-based features can also be used to infer gene expression with tools such as LIQUORICE^33^ and Griffin^34^, which could enable detection of cancer-type and subtype specific signals for diagnosis of patients and, potentially, greater sensitivity. Greater sensitivity could also be achieved by integrating cfDNA fragmentation and methylation into a single machine learning classifier, as the use of machine learning improved sensitivity and specificity of disease detection with cfDNA methylation in this study. Although cfDNA was analysed for four different modalities in our study, there are additional modalities such as single nucleotide variants (SNVs) and fusion events^9–13^ that could further improve a multi-modal liquid biopsy for EwS; for example, detection of *STAG2* mutations may be prognostic^35^. Moreover, other analytes, such as cfRNA may improve sensitivity^36^ and inform on toxicity^37^. Future studies should include additional liquid biopsy analytes, such as cfRNA, as well as exploring further modalities and signals from cfDNA as they could reflect different biological processes.

In conclusion, this study adds to the previous publications on liquid biopsies that begin to show promise for monitoring of EwS patients and detection of relapse events. Further validation is now warranted in larger patient cohorts to determine the optimal combination of blood-based analytes (cfDNA features and other approaches) and their clinical utility embracing a minimally invasive approach to augment imaging for children and young adults with EwS.

## 4 Materials and Methods

### 4.1 Study design and sample collection

Patients with EwS or Ewing-like Sarcoma were recruited under the Manchester Cancer Research Centre (MCRC) Biobank EwS project (application 21_CADI_03) and consented to give samples for this study with appropriate ethical approval from the MCRC Biobank Tissue Bank Ethics (ref: 22/NW/0237). All patients were recruited at The Christie NHS Foundation Trust and the Royal Manchester Children’s Hospital.

Non-cancer control samples were collected from three sources, with informed consent: 1. The Community Lung Health Study (ethically approved study London – West London & GTAC Research Ethics Committee REC reference: 17/LO415); 2. The University of Manchester healthy normal volunteer study (University of Manchester Research Ethics Committee 4 (UREC4) approval no. 2017-2761-4606); or 3. Purchased from Cambridge Bioscience (University of Manchester Research Ethics Committee approval no. 2019-7920-11797).

### 4.2 Blood sample collection

Up to 10 mL of whole blood was collected from patients in Streck DNA blood collection tubes (BCTs). Blood was spun at 2000 x g for 10 minutes at room temperature with the brake off. The plasma supernatant was collected and centrifuged again at 2000 x g for 10 minutes at room temperature. The resulting supernatant was collected, aliquoted and stored at -80°C. Blood was processed after a minimum of 24 hours at room temperature, and within 96 hours of collection.

### 4.3 cfDNA isolation and QC

cfDNA from patients with EwS or CIC were isolated from Streck DNA plasma (median = 4.5 mL, range = 1.5 - 6.5 mL) using the EZ1&2 ccfDNA Kit (QIAGEN) on the EZ2 Connect liquid automation platform (QIAGEN) using a custom protocol which used 8 mL plasma input and 45 μL elution volume. If Streck DNA plasma volume was less than 8 mL, PBS was used to supplement the volume up to 8 mL. For the NCC samples (not previously published), cfDNA was isolated from Streck DNA plasma using the QIAamp MinElute ccfDNA kit (QIAGEN) for all samples except NCC0112, which was isolated using the QIAsymphony DSP Circulating DNA kit (QIAGEN) on the QIAsymphony instrument (QIAGEN). All isolated cfDNA was quantified using the TaqMan™ RNase P Detection Reagents Kit (Applied Biosystems, Catalog number: 4316831) with a standard curve (maximum of 1000 copies, then 2-fold down to minimum of 15.625 copies). cfDNA quality was analysed using the TapeStation 2100 (Agilent) and the Cell-free DNA ScreenTape Analysis (Agilent) according to the manufacturer’s instructions.

One sample (patient 14, day 14) presenting with haemolysis underwent a bead-based size selection to preferentially remove high molecular weight material (>500 bp). Briefly, a 0.5X bead ratio was added to the sample diluted 1:4.4 in low TE buffer (ThermoFisher). The beads were separated from the solution with a magnet, and the eluate containing low molecular weight material was kept. The eluate then underwent a bead-based clean up with a 2.5X bead ratio (AMPure XP Beads, Beckman Coulter) to remove salts that may interfere with downstream reactions.

### 4.4 T7-MBD-seq library preparation

T7-MBD-seq was performed as described by Conway et al.^25^ using cfDNA (input 5.25 – 25 ng). cfDNA samples underwent end-repair and dA-tailing (NEBNext® Ultra™ II End Repair/dA-Tailing Module, New England Biolabs), dephosphorylation (FastAP Thermosensitive Alkaline Phosphatase (1 U/μL), Fisher Scientific), then ligation to a sample-specific barcode overnight (Kapa HyperPrep Kit PCR-Free, Roche and custom oligo, Integrated DNA Technologies). After barcode ligation, samples were pooled (9 to 13 samples per pool, 112 to 254 ng per pool) and remaining adapters removed via 0.8X bead clean-up (AMPure XP Beads, Beckman Coulter). 90% of the sample pool underwent a methylation enrichment with the EpiMark® Methylated DNA Enrichment Kit (New England Biolabs) with the remaining 10% of the library pool retained for WGS. Both the enriched and unenriched sample pools were amplified by HiScribe T7 High Yield RNA Synthesis Kit (New England Biolabs) overnight to produce amplified RNA (aRNA). Methylation enriched aRNA underwent a single 1X bead clean-up and WGS aRNA underwent two 1X bead clean-ups prior to quality control checks using agarose gel electrophoresis and Qubit RNA Broad Range assay (ThermoFisher Scientific). All aRNA was dephosphorylated (FastAP Thermosensitive Alkaline Phosphatase (1 U/uL), Fisher Scientific), then ligated to an adapter containing an Illumina read 2 sequencing primer-compatible sequence (Custom oligonucleotide, Integrated DNA Technologies) overnight. Ligated aRNA underwent a 1X bead clean-up prior to reverse transcription (Superscript IV Reverse Transcriptase, ThermoFisher Scientific). A 1.5X bead clean-up was performed prior to PCR (KAPA HiFi PCR Kit HotStart PCR Kit, with dNTPs 250U, Roche) with unique dual index primers. A final 1.8X bead clean-up was performed prior to library quantification by qPCR (KAPA Library Quantification Kit for Illumina Universal, Roche). The enriched and unenriched library pool were then pooled equimolar, and then together at a ratio of 2:3. This pool was sequenced 150 bp paired-end on an Illumina NovaSeq6000 with S4 reagents (Illumina) and 15% PhiX.

### 4.5 Bioinformatic data analysis software

All data analysis was performed in R (v4.3.0) using RStudio (2024.12.0+467.pro1 “Kousa Dogwood” Release). R packages used are listed in supplementary information.

### 4.6 T7-MBD-seq analysis

T7-MBD-seq analysis was performed as described previously^25^. Briefly, a Nextflow (24.04.2)^38^ DSL2 pipeline^25^ was used for FASTQ processing and generation of QSEA objects. Reads were trimmed to 91/61 basepairs (bp) for read 1 and read 2 respectively. The unique molecular identifier (UMI) was extracted using umitools (v1.1.5)^39^ and samples were demultiplexed and adapters trimmed with cutadapt (v3.4)^40^. Reads were aligned to the reference genome (GRCh38) using bwa mem (0.7.18)^41^, deduplicated using both the R1 start position and UMIs with umi-tools (v1.1.5) and mate quality scores assigned with samtools fixmate (1.16.1)^42^. FastQC (v0.11.9), Qualimap (v2.2.2d) and MultiQC (v1.13) were used throughout the pipeline for quality check. The QSEA package (v4.3.2)^43^ was used to analyse bam files alongside the R package MESA (v0.5.1, from https://github.com/cruk-mi/mesa). In brief, the genome was tiled into 300 bp non-overlapping windows as previously described^25^. Only paired reads where either end of the pair mapped with a Mapping Quality (MAPQ) score of at least 10, had a fragment length between 70 and 1000 bp and a distance along the reference genome of at least 30 bp were used in downstream analysis. Fragments were then uniquely assigned into windows according to the location of their midpoint. Copy Number Variations (CNV) were calculated for each sample from the non-enriched fraction, using hmmcopy (v1.44.0)^44^ with base parameters over 1 Mbp windows. Normalised reads per million (NRPM) were generated using the CNV and the number of valid fragments in the sample, without applying trimmed mean of M values (TMM) normalisation. Beta-values (a scaled measure of methylation between 0 and 1) for each window in each sample were calculated within QSEA using the blind calibration method.

### 4.7 IchorCNA

cfDNA WGS data was processed with ichorCNA (v0.3.2) to give an estimated tumour fraction (ichorTF) using a panel of normal samples. Estimated ichorTF below 0.03 were deemed to be below the limit of detection^27^.

### 4.8 Quality Checks

NGSCheckMate (v1.0.1)^45^ was used to confirm that all longitudinal samples as well as enriched and WGS T7-MBD-seq libraries from the same individual matched as expected. The relative enrichment score (relH) was calculated using the MEDIPs R package^46^ for enriched libraries to confirm enrichment of CpGs, as described by Conway et al.^25^. The hyperstable fraction^25^ was also calculated using consistently hypermethylated CpGs^47^. Adequate methylation capture was deemed to be achieved when the relH was above 3.0 and the hyperstable fraction was above 0.4. All samples used in this analysis were deemed to have adequate methylation capture.

### 4.9 Statistics

For p100-150 bp and β mean, the thresholds for positivity were determined using a T-score cut-off approach. First, the T-score (T) was calculated for the upper tail using the inverse cumulative density function of the Student t distribution (qt function, R) where the vector of probabilities (p) was 0.025 and the degrees of freedom were n-1. The margin of error was then calculated using Tσ_M_. Finally, the cut-off was calculated using μ + margin of error. The cut-off was calculated using either the test set NCCs and on treatment samples, and the highest cut-off was applied (p100-150 bp on treatment cut-off used, β mean NCC cut-off used).

All statistical tests performed are detailed in the text and figures. Where a Wilcoxon test was performed, stat_compare_means was used (ggpubr, v0.6.0) to calculate and add the p-value to each plot. Non-significant p-values are not plotted. Where detail was not provided, all tests were 2-sided with a threshold of p < 0.05.

### 4.10 EwingSign Classifier

An EwS methylation-based classifier, EwingSign, was developed using an approach adapted from Conway et al.^25^ to train a machine learning model capable of predicting three distinct classes: EWS, CIC, or non-cancer. To construct this classifier, we obtained methylation array data for EwS and CIC from the Gene Expression Omnibus (accession number GSE140686^21^), which included 11 CIC arrays (8 from the 450k platform and 3 from the EPIC platform) and 37 EwS arrays (26 from the 450k platform and 11 from the EPIC platform). The following steps were performed within a Nextflow (24.04.2)^38^ DSL2 pipeline. The SeSaMe R package without pOOBAH (p-value with out-of-band array hybridization) masking was used to process the IDAT files to obtain beta-values for each probe. These methylation array beta-values were then converted into a window-based read format compatible with T7-MBD-seq, as exemplified by Conway et al.^25^. This conversion yielded a QSEA object containing estimated read counts for each sample in windows overlapping the array probes, approximating the counts that would have been obtained if T7-MBD-seq had been performed on the same samples.

For selection of features to use in the classifier, DMRs were identified through pairwise comparisons between EwS and the NCC training set, CIC and the NCC training set, and CIC and EwS samples using the QSEA package. DMRs were selected based on a false discovery rate (FDR) threshold of 0.05. To refine the selection, regions where the median beta value exceeded 0.25 across NCC training set samples were removed from the DMR list. The common DMRs across all comparisons were then identified, and the minimum absolute Δβ (difference in average beta values) was retained for each DMR. Finally, DMRs were ranked in descending order based on Δβ, and the top 200 were selected as features for classifier training, and used in the β mean calculation.

To generate in silico mixture samples, fragment counts were mixed either by incorporating an array sample into an NCC cfDNA training set sample at proportions ranging from 0.005 and 0.1, or by combining two NCC cfDNA training set samples at proportions ranging from 0.15 to 0.5. In total, there were 2200 CIC mixed with NCC, 7400 EWS mixed with NCC and 10000 NCC mixtures used. We trained 100 sub-classifiers using Extreme Gradient Boosting Trees (XGBoost, implemented in the R package xgboost) with default parameters, except for setting the number of trees to 200. Each sub-classifier was trained on a different 80:20 split of samples from the in-silico mixture data, using the same top 200 DMRs described above as features. An ensemble classifier score was then calculated using the mean prediction score for each class across all 100 sub-classifiers. A sample was determined to be CIC, EWS or NCC when a single class reached a mean prediction score > 0.5. An unseen test set consisted of T7-MBD-seq cfDNA data from 29 NCC samples, 58 EwS samples from 15 patients, and 10 CIC samples from 3 patients. These samples were not included in the training process and were used to independently evaluate the performance of the ensemble classifier.

## Data Availability

Data produced in the present work is available in the manuscript or supplementary tables and figures. Raw and processed T7-MBD-seq data will be deposited in a repository after peer review.

## Resource availability

### Lead contact

Further information and requests for resources and reagents should be directed to and will be fulfilled by the lead contacts, Florent Mouliere and Martin McCabe.

### Materials availability

This study did not generate new, unique reagents.

#### Acknowledgements

This work was funded by the CRUK National Biomarker Centre (CTRNBC-2022/100001), The Christie Charitable Trust and the Children’s Cancer Research Fund (DIVE/2023). Support was received from the Manchester Experimental Cancer Medicines Centre and the National Institute for Health and Care Research (NIHR) Manchester Biomedical Research Centre (BRC) (NIHR203308). The authors thank Ekram Aidaros-Talbot, Lee-Anne van Winkel, Matthew Lancashire, and all the members of the NBC for their constant help and support. The authors thank the Manchester Cancer Research Centre Biobank and Royal Manchester Children’s Hospital for assistance with sample collection and processing, and Holly Cassell for training in bioinformatic pipelines. The authors thank the Core Facilities at the Cancer Research UK Manchester Institute (C5759/A27412), with particular thanks to the Molecular Biology Core Facility. The authors also would like to thank the healthy volunteers, patients with EwS and CIC, and their families for consenting to this study.

#### Author Contributions

Conception and design: SAR, MGMC, FM

Experiments and data collection: SAR, GG, AR, AC

Data processing: SAR, PJBH, MK, SMH

Software development: SAR, PJBH, MK, SMH, SPP

Data analysis: SAR, MK, SMH, FM

Sample acquisition: BB, ATJL, MGM

Funding acquisition: CD, DGR, MGM, FM

Manuscript draft: SAR, MK, FM

Manuscript revisions and comments: SAR, GG, AR, PJBH, MK, SMH, SPP, VJM, BB, ATJL, AC, CD, DGR, MGM, FM

Supervision: CD, DGR, MGM, FM

#### Declaration of Interests

S.M.H, S.P.P., A.C., C.D., D.G.R and F.M. are co-inventors on patents related to cell-free DNA analysis. F.M. has consulted for Roche Dx. A.C and F.M have received support from Biomodal. C.D. has received research funding/educational research grants from Amgen, AstraZeneca, Astex Pharmaceuticals, Angle PLC, Boehringer Ingelheim, Carrick Therapeutics, Celgene, Clearbridge Biomedics, CV6 Therapeutics (NI) Ltd, Epigene Therapeutics Inc, GSK, Guardant, Menarini, Merck AG, Neomed Therapeutics, Novartis, RedX Pharma, Roche, Taiho Oncology, Thermo Fisher Scientific, UCB Pharma. C.D. has received honoraria for consultancy and/or advisory boards from Biocartis, Merck, AstraZeneca, GRAIL, Boehringer Ingelheim. A.T.J.L. has received honoraria and support with conference attendance from Alexion Pharmaceutical. All other authors declare no competing interests.

---

**Supplementary Figure 1.**
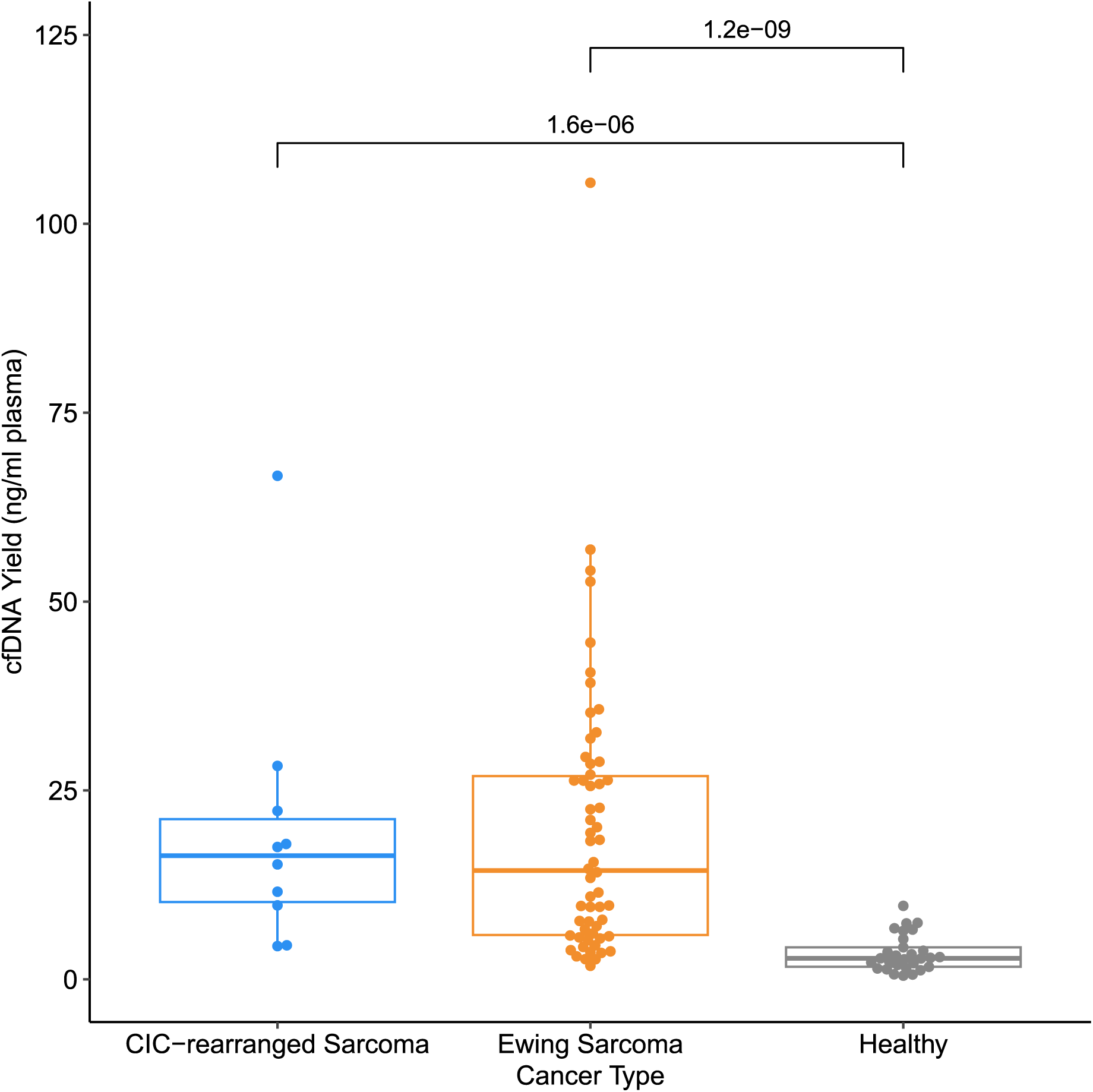
cfDNA yields were greater in CIC and EwS compared to NCCs. cfDNA yields in ng per mL of plasma for all CIC (blue, n = 10 samples), EwS (orange, n = 58 samples) and NCC (grey, n = 29 samples). For the boxplots, the central line indicates the median, and the lower and upper boundaries of the box correspond to the 25^th^ and 75^th^ quartile respectively. The upper whisker extends to the largest value no further than 1.5 * the interquartile range (IQR), and the lower whisker to the smallest value no further than 1.5 * IQR. Each point indicates an individual sample. Wilcoxon test used for comparisons, only significant p values shown (p < 0.05).

**Supplementary Figure 2.**
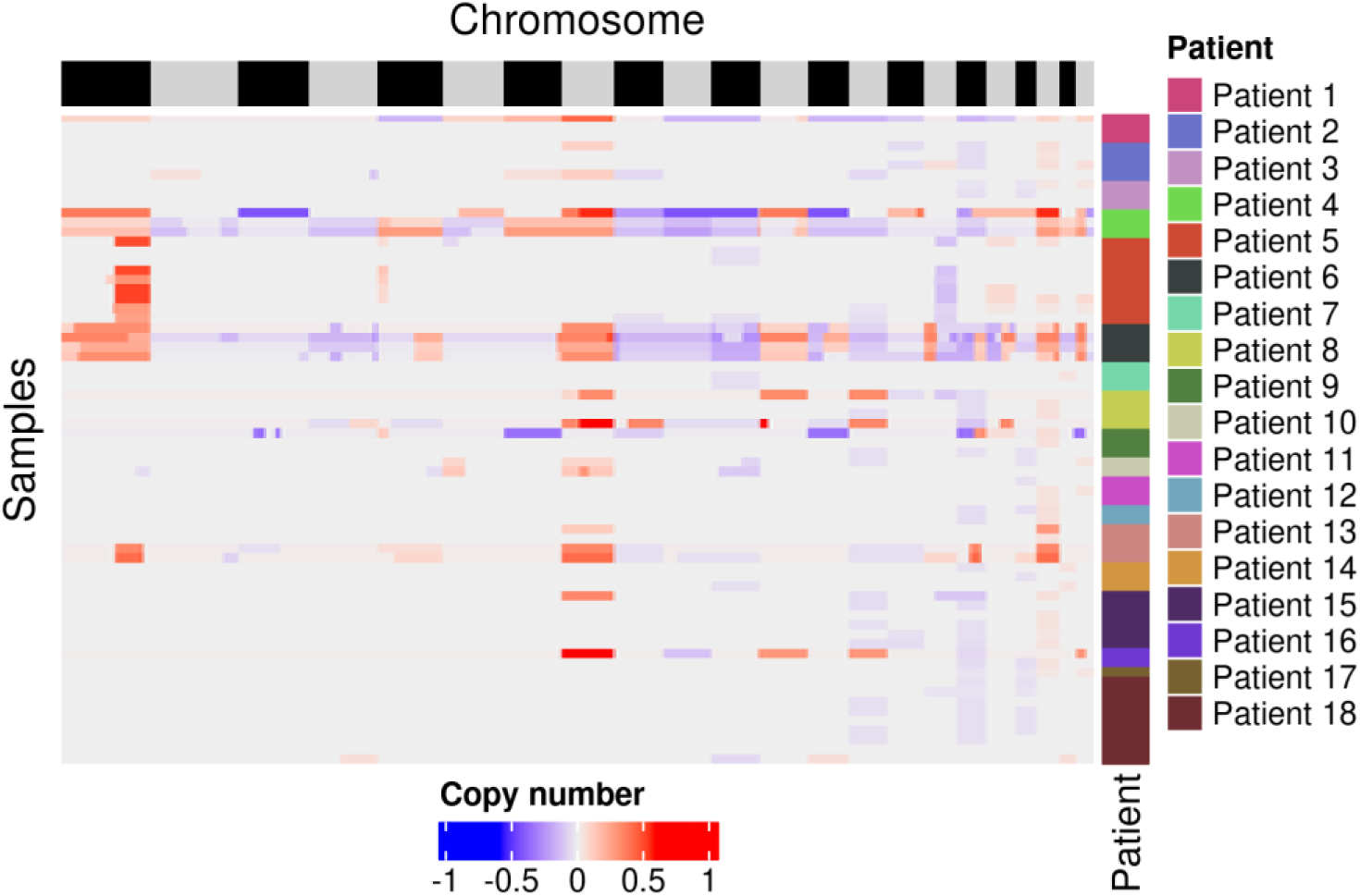
Copy number aberrations in EwS and CIC. Heatmap of CNAs. Each row is a sample with patient annotated on the right. Each column is a region of the genome (1,000,000 bp windows) and separate chromosomes denoted by the top annotation. Gains = red. Losses = blue.

**Supplementary Figure 3.**
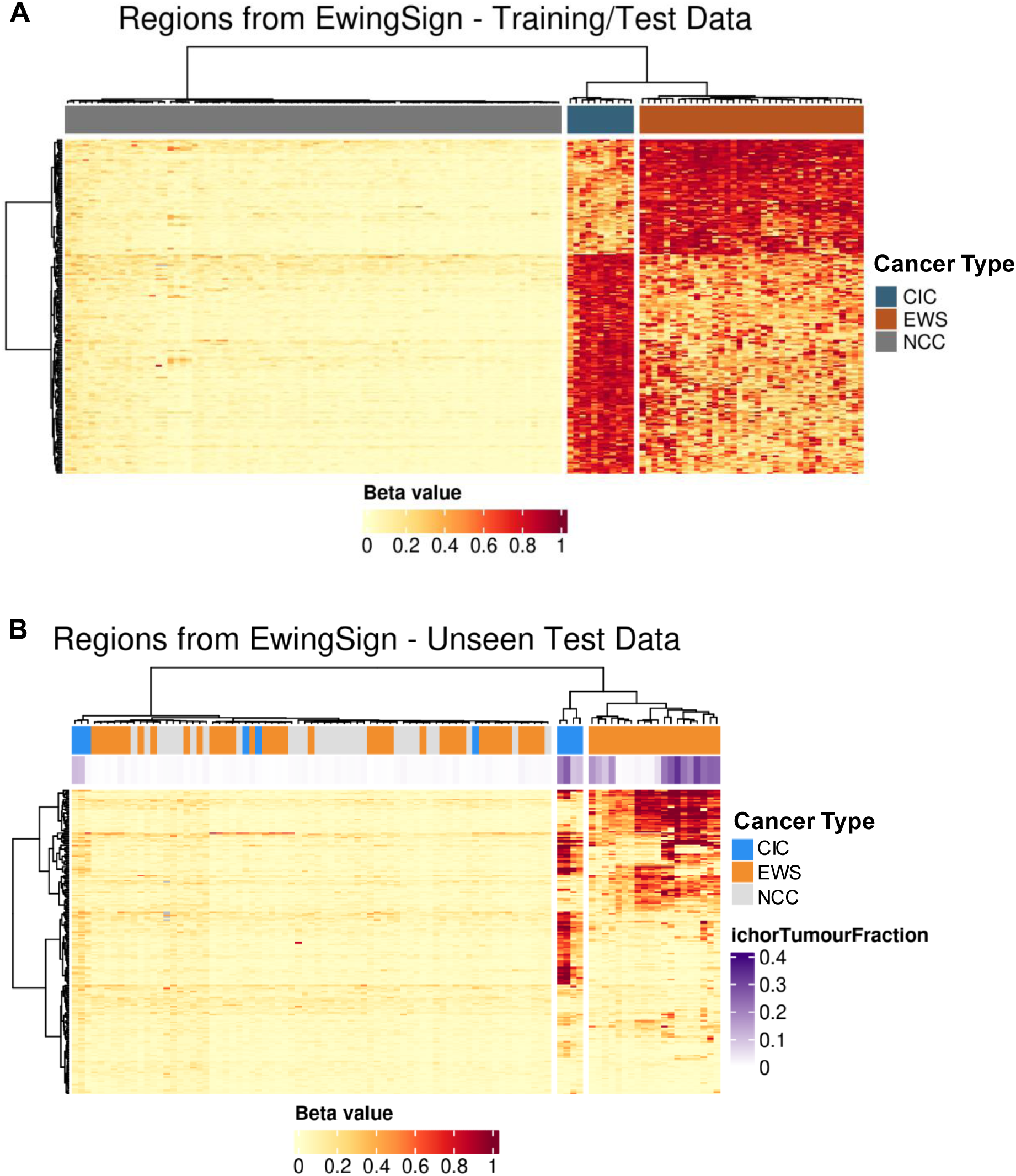
200 differentially methylated regions (DMRs) used to calculate β mean and as features in EwingSign. Heatmap of 200 DMRs used in β mean and EwingSign applied to the training and test data (A) and the unseen test set (B). Each column is a sample, and each row is a DMR. Top annotations indicate the cancer type of the sample (blue = CIC, orange = EWS, grey = NCC) and ichorTF (ichorTF = 0, white; ichorTF > 0.4, purple) (B). High methylation (β = 1) is indicated by red, and low methylation (β = 0) by yellow.

**Supplementary Figure 4.**
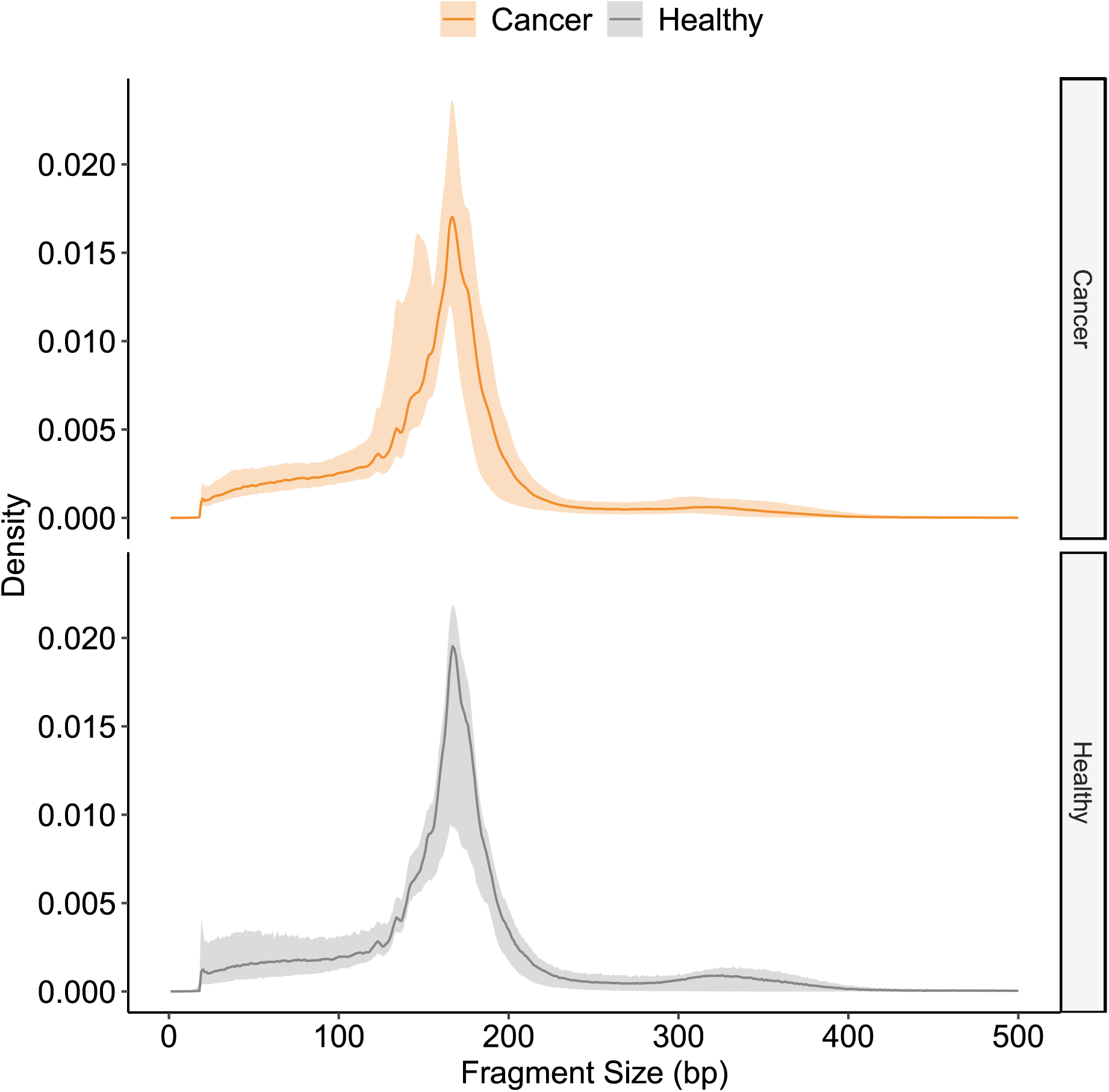
cfDNA fragment size distribution for EwS and CIC, and NCC. The line indicates the median size distribution for that group, and the ribbon indicates the range. EwS and CIC (orange); NCC (grey).

**Supplementary Figure 5.**
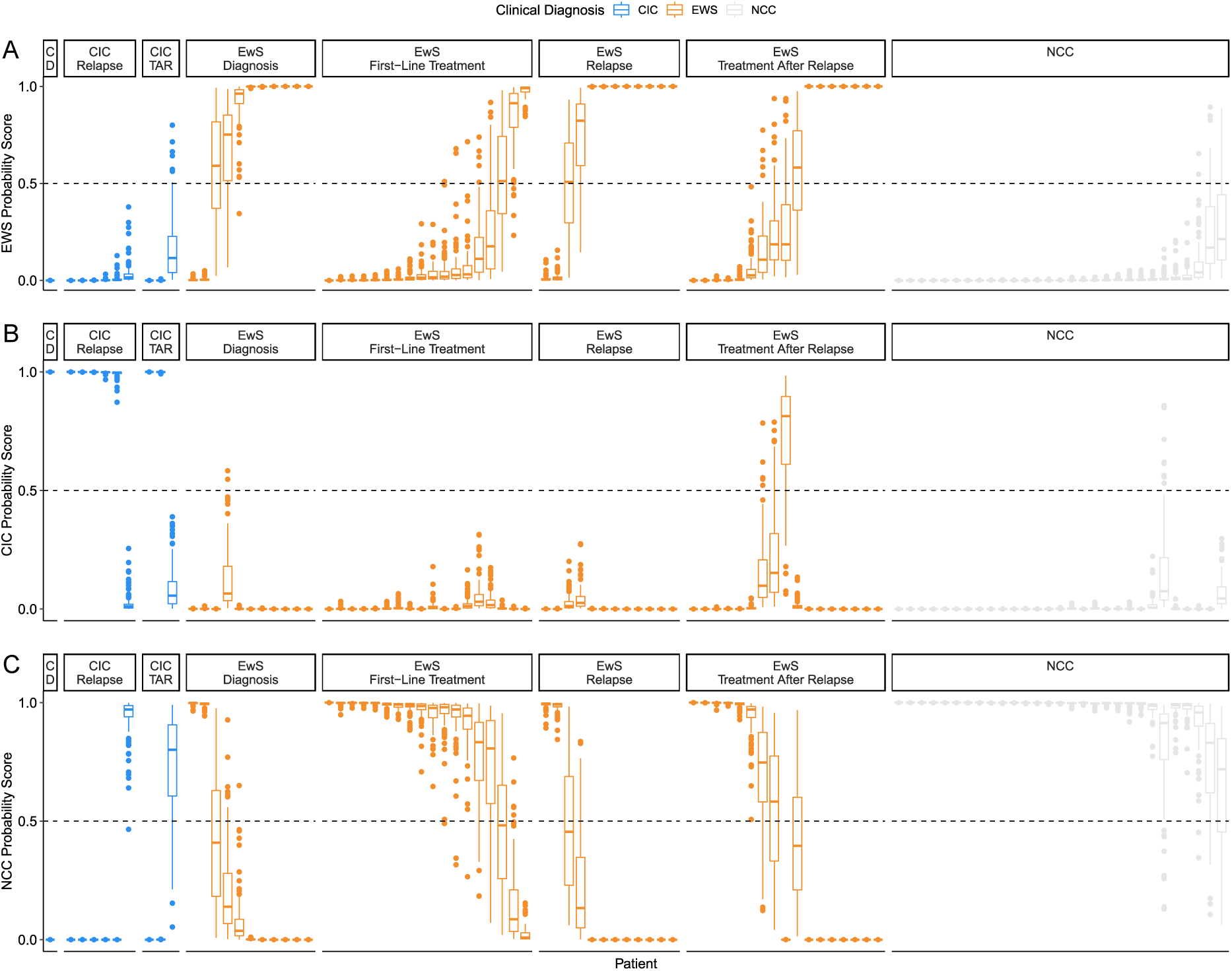
Results from all 100 iterations of the EwingSign sub-classifiers. **A-C:** Boxplots showing the results of all 100 sub-classifiers of EwingSign as EwS score (A), CIC score (B) and NCC score (C). The colour of the boxplot indicates the clinical diagnosis of the sample (CIC = blue, EwS = orange, NCC = grey), with outliers shown as points. All plots are faceted by clinical diagnosis and the timepoint the sample was taken. Facet labels are abbreviated where necessary: C = CIC, D = Diagnosis, TAR = Treatment after relapse.

**Supplementary Figure 6.**
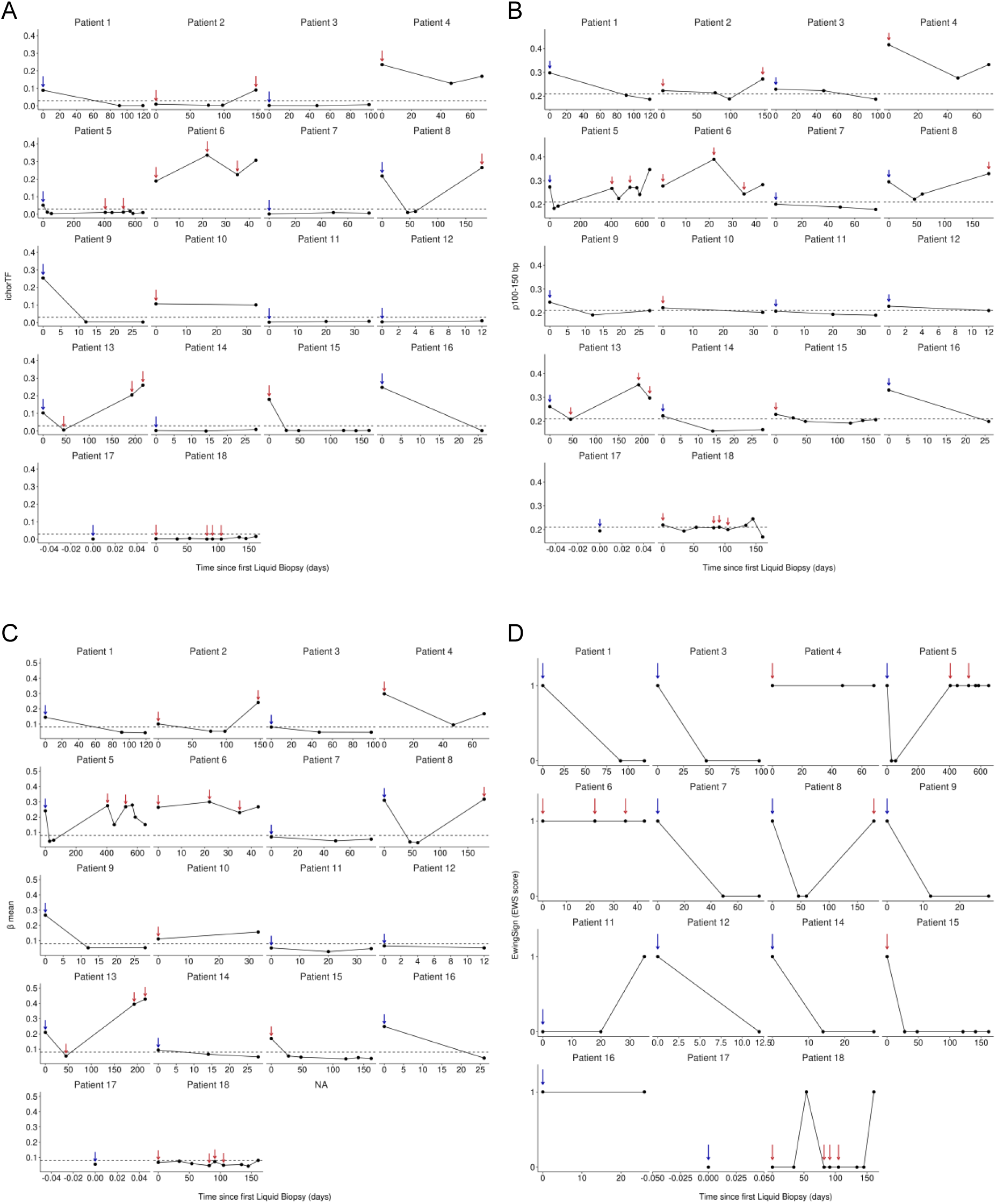
Longitudinal liquid biopsy monitoring of patients with EwS and CIC. **A-D:** Longitudinal monitoring over time (days) using ichorTF (**A**), p100-150 bp (**B**) and β mean (**C**) for all patients. EwingSign (1 = positive for EwS, 0 = negative for EwS, **D**) is shown only for patients with EwS. Blue arrows indicate diagnosis events, red arrows indicate relapse events, and an absence of arrows indicates an on-treatment event. Each point indicates a liquid biopsy sample taken, and its result.

**Supplementary Table 1.** Assay performance for all modalities independently and in combination for detection of disease inclusive of diagnosis and relapse timepoints. PPV – Positive Predictive Value, NPV – Negative Predictive Value.

| **Modality** | **Detected**<br>**Diagnosis** | **Detected**<br>**Relapse** | **Detected**<br>**NCC** | **Sensitivity** | **Specificity** | **PPV** | **NPV** | **Accuracy** |
| --- | --- | --- | --- | --- | --- | --- | --- | --- |
| **ichorTF** | 6/12 | 10/18 | 0/29 | 53.33% | 100% | 100% | 51.72% | 68.89% |
| **p100-150 bp** | 9/12 | 15/18 | 2/29 | 80.00% | 93.10% | 92.31% | 81.82% | 86.44% |
| **β mean** | 9/12 | 14/18 | 6/29 | 76.67% | 79.31% | 79.31% | 76.67% | 77.97% |
| **EwingSign** | 10/12 | 15/18 | 0/29 | 83.33% | 100% | 100% | 85.29% | 91.53% |
| **p100-150 bp +**<br>**EwingSign** | 10/12 | 15/18 | 2/29 | 83.33% | 93.10% | 92.59% | 84.38% | 88.14% |
| **ichorTF + p100-150**<br>**bp** | 9/12 | 15/18 | 2/29 | 80.00% | 93.10% | 92.31% | 81.82% | 86.44% |
| **ichorTF + EwingSign** | 10/12 | 15/18 | 0/29 | 83.33% | 100% | 100% | 85.29% | 91.53% |
| **ichorTF + p100-150**<br>**bp + EwingSign** | 10/12 | 15/18 | 2/29 | 83.33% | 93.10% | 92.59% | 84.38% | 88.14% |

## Notes

### Author Declarations

North West - Greater Manchester South Research Ethics Committee of NHS Health Research Authority gave ethical approval for this work via the Manchester Cancer Research Centre Biobank (22/NW/0237). Non-cancer control samples were collected from three sources, with informed consent: 1. The Community Lung Health Study (ethically approved study London - West London & GTAC Research Ethics Committee REC reference: 17/LO415); 2. The University of Manchester healthy normal volunteer study (University of Manchester Research Ethics Committee 4 (UREC4) approval no. 2017-2761-4606); or 3. Purchased from Cambridge Bioscience (University of Manchester Research Ethics Committee approval no. 2019-7920-11797).

