## Supplementary Information for "Cell-free DNA methylome and fragmentome analysis for disease relapse monitoring in patients with Ewing Sarcoma"

R version 4.3.0 (2023-04-21)

Platform: x86_64-pc-linux-gnu (64-bit)

Running under: Rocky Linux 8.7 (Green Obsidian)

attached base packages:

[1] grid	stats	graphics grDevices utils	datasets methods	base

other attached packages:

[1] nbctools_1.0.1	gridExtra_2.3	cowplot_1.1.3

[4] caret_7.0-1	lattice_0.22-7	ggforce_0.4.2

[7] qs_0.27.3	ggbeeswarm_0.7.2	circlize_0.4.16

[10] patchwork_1.3.0	ggh4x_0.3.0	swimplot_1.2.0

[13] UpSetR_1.4.0	ComplexHeatmap_2.16.0 ggvenn_0.1.10

[16] ggpubr_0.6.0	rstatix_0.7.2	mesa_0.5.1

[19] qsea_1.26.0	lubridate_1.9.4	forcats_1.0.0

[22] stringr_1.5.1	dplyr_1.1.4	purrr_1.0.4

[25] readr_2.1.5	tidyr_1.3.1	tibble_3.2.1

[28] ggplot2_3.5.1	tidyverse_2.0.0

loaded via a namespace (and not attached):

[1] splines_4.3.0	BiocIO_1.10.0

[3] bitops_1.0-9	polyclip_1.10-7

[5] janitor_2.2.1	hardhat_1.4.1

[7] pROC_1.18.5	XML_3.99-0.18

[9] rpart_4.1.24	lifecycle_1.0.4

[11] doParallel_1.0.17	globals_0.16.3

[13] vroom_1.6.5	MASS_7.3-58.4

[15] backports_1.5.0	magrittr_2.0.3

[17] limma_3.56.2	rmarkdown_2.29

[19] yaml_2.3.10	RColorBrewer_1.1-3

[21] abind_1.4-8	zlibbioc_1.46.0

[23] GenomicRanges_1.52.1	BiocGenerics_0.46.0

[25] RCurl_1.98-1.17	nnet_7.3-20

[27] tweenr_2.0.3	ipred_0.9-15

[29] lava_1.8.1	GenomeInfoDbData_1.2.10

[31] IRanges_2.34.1	S4Vectors_0.38.2

[33] listenv_0.9.1	parallelly_1.43.0

[35] codetools_0.2-20	DelayedArray_0.26.7

[37] RApiSerialize_0.1.4	tidyselect_1.2.1

[39] shape_1.4.6.1	HMMcopy_1.42.0

[41] farver_2.1.2	matrixStats_1.5.0

[43] stats4_4.3.0	GenomicAlignments_1.36.0

[45] GetoptLong_1.0.5	e1071_1.7-16

[47] Formula_1.2-5	survival_3.8-3

[49] iterators_1.0.14	systemfonts_1.2.1

[51] foreach_1.5.2	tools_4.3.0

[53] ragg_1.3.3	Rcpp_1.0.14

[55] glue_1.8.0	prodlim_2024.06.25

[57] xfun_0.52	MatrixGenerics_1.12.3

[59] GenomeInfoDb_1.36.4	withr_3.0.2

[61] fastmap_1.2.0	digest_0.6.37

[63] timechange_0.3.0	R6_2.6.1

[65] textshaping_1.0.0	colorspace_2.1-1

[67] Cairo_1.6-2	gtools_3.9.5

[69] generics_0.1.3	data.table_1.14.8

[71] recipes_1.2.1	rtracklayer_1.60.1

[73] class_7.3-23	S4Arrays_1.2.0

[75] ModelMetrics_1.2.2.2	pkgconfig_2.0.3

[77] gtable_0.3.6	timeDate_4041.110

[79] rsconnect_1.3.4	XVector_0.40.0

[81] htmltools_0.5.8.1	carData_3.0-5

[83] plyranges_1.20.0	clue_0.3-66

[85] scales_1.3.0	Biobase_2.60.0

[87] hues_0.2.0	png_0.1-8

[89] snakecase_0.11.1	gower_1.0.2

[91] knitr_1.50	rstudioapi_0.17.1

[93] tzdb_0.5.0	reshape2_1.4.4

[95] rjson_0.2.23	nlme_3.1-168

[97] proxy_0.4-27	cachem_1.1.0

[99] zoo_1.8-13	GlobalOptions_0.1.2

[101] parallel_4.3.0	vipor_0.4.7

[103] restfulr_0.0.15	pillar_1.10.1

[105] vctrs_0.6.5	car_3.1-3

[107] stringfish_0.16.0	cluster_2.1.8.1

[109] beeswarm_0.4.0	evaluate_1.0.3

[111] magick_2.8.6	cli_3.6.4

[113] compiler_4.3.0	Rsamtools_2.16.0

[115] rlang_1.1.5	crayon_1.5.3

[117] future.apply_1.11.3	ggsignif_0.6.4

[119] labeling_0.4.3	plyr_1.8.9

[121] stringi_1.8.7	BiocParallel_1.34.2

[123] munsell_0.5.1	Biostrings_2.68.1

[125] Matrix_1.6-5	BSgenome_1.68.0

[127] hms_1.1.3	bit64_4.6.0-1

[129] future_1.34.0	SummarizedExperiment_1.30.2

[131] broom_1.0.8	memoise_2.0.1

[133] RcppParallel_5.1.10	bit_4.6.0
